# Impact of age, gender, ethnicity and prior disease status on immunogenicity following administration of a single dose of the BNT162b2 mRNA Covid-19 Vaccine: real-world evidence from Israeli healthcare workers, December-January 2020

**DOI:** 10.1101/2021.01.27.21250567

**Authors:** Kamal Abu Jabal, Hila Ben-Amram, Karine Beiruti, Yunis Batheesh, Christian Sussan, Salman Zarka, Michael Edelstein

## Abstract

The Pfizer Covid-19 vaccine showed high efficacy in clinical trials but observational data from populations not included in trials are needed. We described immunogenicity 21 days post-dose 1 among 514 Israeli healthcare workers by age, gender, ethnicity and prior COVID19 infection. Immunogenicity was similar by gender and ethnicity but decreased with age. Those with prior infection had antibody titres one magnitude order higher than naïve individuals regardless of the presence of detectable IgG antibodies pre-vaccination.

## Background

Despite multiple lockdowns and prolonged control measures implemented in most countries the COVID19 pandemic, which started in 2019 in China, continues to spread. As of January 25 2021, over 99 million cases and 2.1 million deaths were reported globally[1]. In December 2020, several vaccine candidates were shown to be safe and effective [2-4] and mass vaccination (in combination with existing control measures) is seen as the most likely solution to end the pandemic. Although clinical trial data are encouraging, real world evidence with regards to the vaccines remains scarce. In particular, describing immunogenicity and efficacy among specific ethnic groups is important as the virus disproportionately affects certain ethnic minorities for reasons not fully understood but not fully attributable to socio-demographic factors [5-6]. Likewise, understanding the added value of vaccination among those who were infected remains an unanswered question.

As of 25 January 2021, Israel had vaccinated 29.2 % of its population with a single dose of vaccine [7], the highest proportion in the world, almost exclusively with the Pfizer BNT162b2 mRNA Covid-19 Vaccine. Healthcare workers were among the first to be eligible. Ziv Medical Center (ZMC)), located in Safed, Israel, is a large hospital serving the North of Israel. It is staffed by a multi-ethnic workforce of approximately 1500 including Jews, Arabs and Druze among others. Starting December 2020 ZMC has offered the mRNA based Pfizer vaccine to all its staff, and as of 21 January 2021, 1-dose uptake was approximately 90%. In order to describe 1-dose immunogenicity among different groups we measured antibody levels following administration of one dose of the Pfizer vaccine and reported them according to age, gender, ethnicity and prior disease status (measured by baseline IgG levels and/or evidence of a previous positive PCR test)

### Measuring COVID-19 antibody levels before and after vaccination

Prior to vaccination, Healthcare workers were consented to their Neutralizing IgG antibody level being measured, using the Abbott Architect SARS-CoV-2 IgG assay, detecting N antibodies with high sensitivity and specificity [8]. Each positive sample was re-tested for verification purposes, using the quantitative LIAISON Diasorin SARS-CoV-2 S1/S2 IgG assay [8]. In order to ascertain prior infection with SARS-CoV-2, we searched for results of any PCR tests for all participants. Workers were also asked to consent to IgG antibody level testing 21 days after dose 1 (at the same time as receiving dose 2), using the quantitative LIAISON Diasorin SARS-CoV-2 S1/S2 IgG assay [8]. The vaccine was offered to all workers regardless of consenting to antibody level measurement. Antibody levels were reported using geometric means concentration alongside 95% confidence intervals (95%CI), stratified by age, gender, ethnicity and prior disease status. We compared the age and ethnicity of non-responder to other using Kurskall Wallis and Chi-square tests, respectively. In order to describe the range of antibody levels we also described the distribution of IgG antibody levels using boxplots of the log10 IgG titres. The study was approved by ZMC’s ethics committee (0133-20-ZIV).

### Impact of gender, age, ethnicity and prior infection on immunogenicity

Of the 1378 workers who received the first dose of the vaccine, 514 (37%) took part in the study and had their antibody levels measured at 21 days. Among those who received the vaccine, 385 (74.9%) were tested for IgG levels at baseline, prior to vaccination. Of these 6 were IgG positive (of which 4 had evidence of a previously positive PCR test); an additional 11 had recorded evidence of a positive PCR test between March and November 2020 but were IgG negative. Among all vaccinated healthcare workers, 475 (92%) had detectable Anti SARS-CoV-2 Spike IgG antibodies and among these geometric means concentration (GMC) was 68.6 AU/ml (95%CI 64-73.6). Those who did not respond to the first dose were older (median age 57 vs 45 in other, p<0.001) and more likely to be Jewish (82% of non-responders vs 63% of responders, p=0.01). Among responders, There was no statistically significant difference in antibody titres between males and females and between different ethnicities, but titres decreased with increasing age (Table 1 and Figure 1). Compared with workers with no evidence of previous disease, post-vaccination IgG levels among those with previous evidence of disease (either a positive IgG at baseline or a previously recorded positive PCR test) were much higher (GMC 573 vs 61.5). IgG titers among those with previous evidence of infection were at least one order of magnitude high than those without, regardless of whether IgG antibodies were detectable prior to being vaccinated (Figure 1).

**Table 1.** Geometric mean concentration of Anti SARS-CoV-2 Spike IgG antibodies among healthcare workers 21 days post 1 dose of the Pfizer/BioIntech COVID19 vaccine

|  |  |  |  |  |  |
| --- | --- | --- | --- | --- | --- |
| Group |  | Number of individuals |  | IgG Geometric means concentration (AU/ml) | 95% CI |
|  |  | N | % |  |  |
| Age | <30 | 11 | 2.1 | 100.4 | 51.8-194.5 |
|  | 30-39 | 161 | 31.3 | 84.2 | 74.3-95.3 |
|  | 40-49 | 146 | 28.4 | 68.2 | 60.2-77.4 |
|  | 50-59 | 101 | 9.7 | 61.5 | 52.6-71.9 |
|  | 60+ | 95 | 18.5 | 49.8 | 42.6-58.1 |
| Ethnicity | Jewish | 322 | 62.6 | 62.4 | 58.2-66.9 |
|  | Arab | 114 | 22.2 | 69.9 | 59.6-82 |
|  | Druze | 58 | 11.3 | 73.4 | 58.6-92 |
|  | Circassian | 2 | 0.4 | - | - |
| Gender | Male | 193 | 37.5 | 64.6 | 60.2-69.2 |
|  | Female | 321 | 62.5 | 75.9 | 65.6-87.9 |
| Prior Disease Status* | All patient with evidence of prior COVID-19 infection | 17 | 3.3 | 573.6 | 289-1138.7 |
|  | IgG positive at baseline | 6 | 1.2 | 747.3 | 140-3978.3 |
|  | IgG negative with prior positive PCR test | 11 | 2.1 | 496.5 | 217.4-1134 |
|  | IgG negative at baseline and no prior positive PCR test | 369 | 71.8 | 61.5 | 58-65.1 |
|  | Unknown (no PCR test and not tested at baseline) | 128 | 24.9 | 64.3 | 60.5-68.3 |

**Figure 1.**
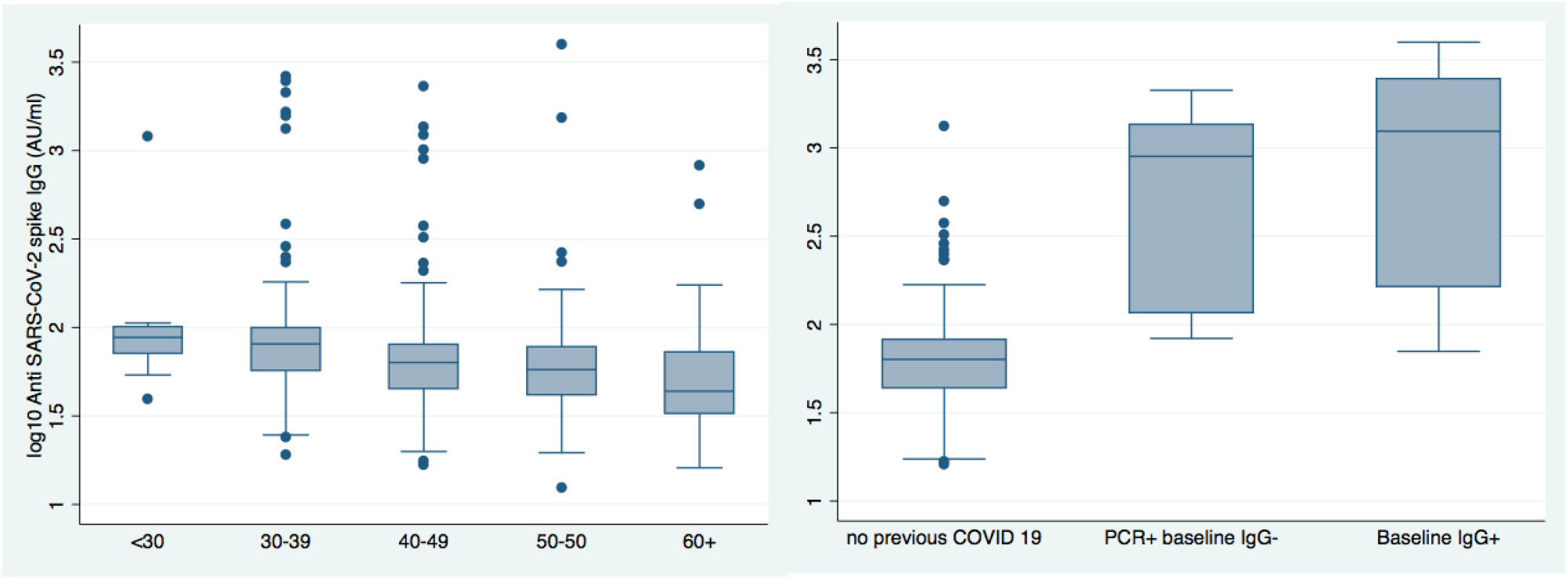
Anti SARS-CoV-2 spike IgG antibody levels among ZMC workers, 21 days post vaccination with 1 dose of the Pfizer/BioIntech COVID19 vaccine by age (left) and prior disease status (right)

## Discussion

A single dose of the Pfizer/BioIntech COVID19 vaccine was immunogenic in the vast majority of our study cohort 21 days post vaccination, a result compatible with trial data [9]. Although the numbers are small, our data however suggests that age and ethnicity may be associated with the likelihood of non-response. Larger scale data should be analysed to confirm or refute such a possibility. There was no significant difference in immunogenicity by gender or ethnicity and although IgG titers were decreasing with increasing age, the differences are small and of doubtful clinical significance in the absence of known correlates of protection. Efficacy data suggests similar efficacy among different age groups [3]. Unsurprisingly, vaccinating individuals with evidence of prior COVID19 infection leads to a boost response, achieving IgG titres at least one order of magnitude of magnitude higher compared with naïve individuals. Interestingly, this was the case in our cohort regardless of whether Anti SARS-CoV2 Nabs were detectable or not immediately prior to vaccination. Although these results are based on small number, these findings provide reassurance that the well documented rapid waning of nucleocapsid IgG Antibodies post acute-COVID19 infection [10] does not translate to a loss of immunity, and the boost-like response seen among previously infected individuals in our cohort suggests B-cell mediated memory immunity is preserved regardless of IgG status. Our study confirms recently published evidence suggesting that immune memory persists at least 6 months post infection [11]. One case in our cohort who showed a boost-type response almost 10 months after testing positive by PCR suggests this could be longer. In situations of scarce vaccine availability it may therefore be safe to assume that most individuals with prior evidence of infection are not prioritized for vaccination, regardless of Anti-Nucletotide IgG levels. Nevertheless, effectiveness of infection against future infection is not 100% protective [12] and offering vaccination to these individuals may confer additional protection, as major Public health agencies recommend [13]. A single dose of vaccine in these individuals seems to boost the response although the optimal timing between infection and vaccination as well as the ensuing duration of protection remain to be determined. Our study only contains a small number of previously infected individuals, as the Israeli Ministry of Health guidelines recommended that these individuals are not prioritized for vaccination, and findings from this study should be replicated on a larger scale in order to make policy decisions. As the immunization programme continues to expand in Israel, previously infected healthcare workers will be offered the vaccine and we will continue to assess Antibody levels in these workers as well as in all others following the administration of the second dose of vaccine. Trends in antibody response following 2 doses of vaccine among the different groups of healthcare workers that comprise ZMC’s workforce are also being analysed and will be shared as they become available.

## Data Availability

The datasets generated during and/or analysed during the current study are available from the corresponding author on reasonable request and with permission from Ziv Medical Centre

